# From Biobanking to Personalized Medicine: the journey of the Estonian Biobank

**DOI:** 10.1101/2024.09.22.24313964

**Authors:** Lili Milani, Maris Alver, Sven Laur, Sulev Reisberg, Toomas Haller, Oliver Aasmets, Erik Abner, Helene Alavere, Annely Allik, Tarmo Annilo, Krista Fischer, Georgi Hudjashov, Maarja Jõeloo, Mart Kals, Liis Karo-Astover, Silva Kasela, Anastassia Kolde, Kristi Krebs, Kertu Liis Krigul, Jaanika Kronberg, Karoliina Kruusmaa, Viktorija Kukuškina, Kadri Kõiv, Kelli Lehto, Liis Leitsalu, Sirje Lind, Laura Birgit Luitva, Kristi Läll, Kreete Lüll, Kristjan Metsalu, Mait Metspalu, René Mõttus, Mari Nelis, Tiit Nikopensius, Miriam Nurm, Margit Nõukas, Marek Oja, Elin Org, Marili Palover, Priit Palta, Vasili Pankratov, Kateryna Pantiukh, Natalia Pervjakova, Natàlia Pujol-Gualdo, Anu Reigo, Ene Reimann, Steven Smit, Diana Sokurova, Nele Taba, Harry-Anton Talvik, Maris Teder-Laving, Neeme Tõnisson, Mariliis Vaht, Uku Vainik, Urmo Võsa, Tõnu Esko, Raivo Kolde, Reedik Mägi, Jaak Vilo, Triin Laisk, Andres Metspalu

## Abstract

Large biobanks have set a new standard for research and innovation in human genomics and implementation of personalised medicine. The Estonian Biobank was founded a quarter of a century ago, and its biological specimens, clinical, health, omics, and lifestyle data have been included in over 800 publications to date. What makes the biobank unique internationally is its translational focus, with active efforts to conduct clinical studies based on genetic findings, and to explore the effects of return of results on participants. In this review we provide an overview of the Estonian Biobank, highlight its strengths for studying the effects of genetic variation and quantitative phenotypes on health-related traits, development of methods and frameworks for bringing genomics into the clinic, and its role as a driving force for implementing personalized medicine on a national level and beyond.

## Background

Large population-based biobanks have set a new standard for the recruitment of individuals, collection and use of biological specimens, data management and harmonisation of medical, genomic, behavioural and lifestyle data for research and innovation. The Estonian Biobank (EstBB), which was established 25 years ago as an independent foundation and later joined the University of Tartu, has placed itself among the world’s largest biobanks. To date, EstBB data has been included in over 300 research projects and over 800 publications. Here, we provide an updated overview of EstBB, highlight its unique strengths for studying the effects of genetic variation and quantitative phenotypes on health-related traits, and its role as a driving force for implementing personalized medicine on a national level and beyond.

Over the past two decades, EstBB has passed through several phases of growth (**Figure 1A**). The first decade focused on participant recruitment and data collection using recruitment personnel, mostly general practitioners (GPs), and special recruitment offices managed by the biobank across the country. In total, 450 GPs and approximately 15 recruitment offices were involved, resulting in a cohort of 52,000 participants by 2015. All individuals signed a broad consent, donated blood samples and were interviewed based on a questionnaire consisting of approximately 330 questions on lifestyle, education, medical history, medication use and female health. The broad consent allows EstBB to regularly update participants’ records by retrieving data from the national health databases of the Estonian Health Insurance Fund (EHIF) and the National Health Information System (NHIS). EHIF manages national health insurance, providing access to healthcare services, medicines, medical equipment, etc., making them the main processor of health insurance data for the entire population. NHIS is the cornerstone of the Estonian e-Health ecosystem, which develops and manages e-services and is thus a junction of health-related information. The questionnaires and linkage to electronic health records (EHRs) provide more than 1,000 data fields covering lifestyle, diet, clinical diagnoses, medication use, and medical procedures^1,2^.

**Figure 1.**
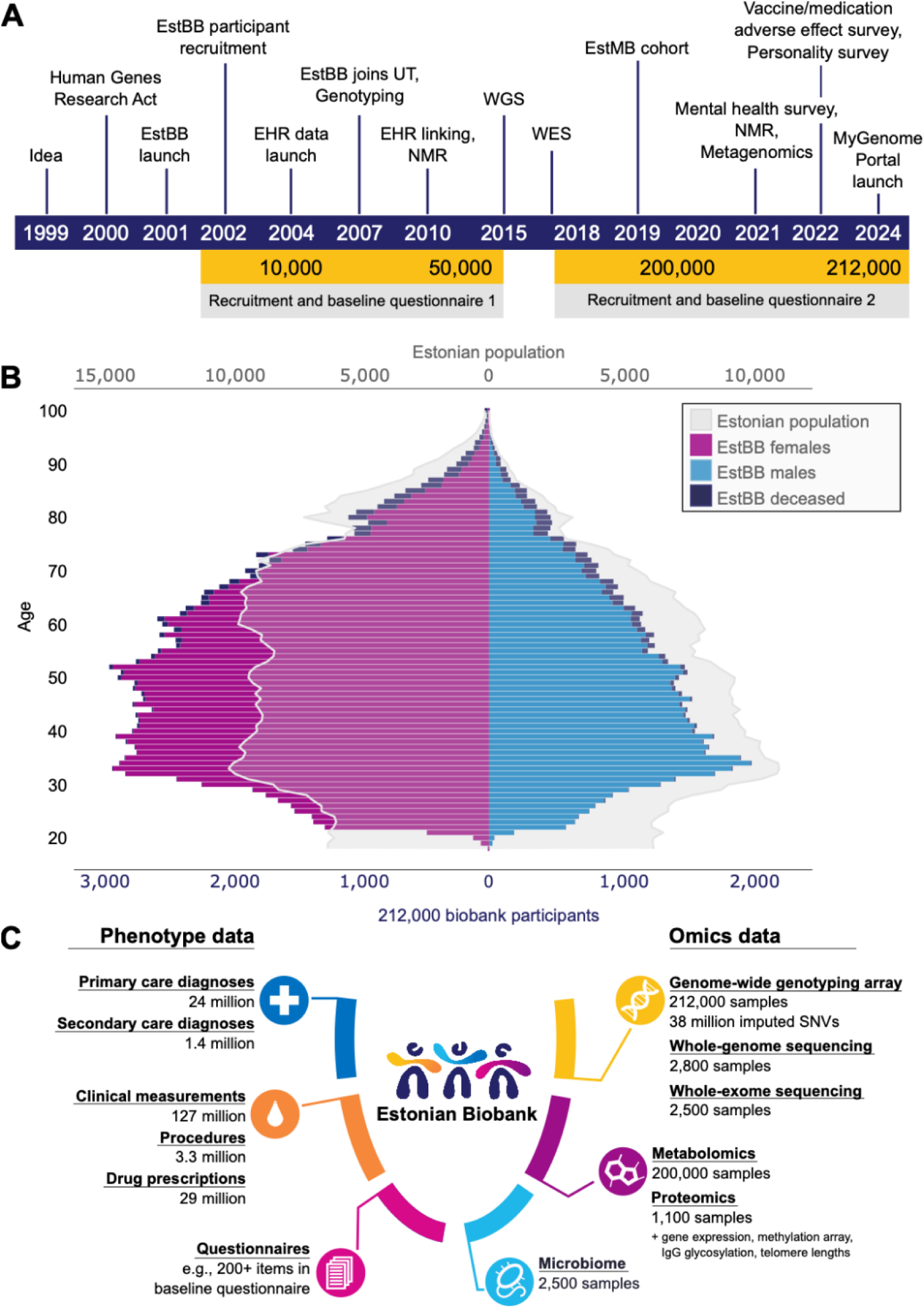
A) Timeline depicting major milestones in EstBB; B) Overview of the age and sex distribution of EstBB participants and comparison to the whole Estonian population in 2023. Different coloured bars correspond to male, female, and deceased participants (blue, purple, and navy, respectively) in each age category, while the grey outline corresponds to the age and sex distribution in the whole Estonian population; C) Overview of different phenotype and omics datasets available in EstBB.

The second phase of EstBB began with genotyping the DNA samples, comparing the genetic structure of the Estonian population to other European populations^3^, and joining international consortia for genome-wide association studies (GWAS)^4–13^. While EstBB has already been part of hundreds of GWAS, contributing to a range of discoveries in human genetics^14–16^, the most striking results based on EstBB data have been in recall by genotype (RbG) studies, a feature relatively unique among the world’s leading biobanks. For instance, EstBB data has been used to evaluate the clinical characteristics of individuals with specific copy number variants (CNV) in their genome^17^, marking the first RbG study in EstBB. Additionally, the ability to re-contact biobank participants with specific genetic profiles has allowed medical experts to join the efforts of evaluating the efficacy of this approach for identifying individuals at elevated risk for hereditary breast and ovarian cancer^18^ or familial hypercholesterolemia^19^.

These studies, combined with research results on polygenic risk scores (PRS)^20,21^, and the translation of existing genotype data into pharmacogenetic recommendations^22^, provided sufficient evidence of the value of genomics in healthcare for the Estonian Government to fund the recruitment and genotyping of an additional 100,000 biobank participants in 2018, and a further 50,000 in 2019. This third stage of expansion was considerably simplified by the broad adoption of digital services in Estonia, e.g., participation only required signing an electronic consent form using the national digital signature scheme (https://www.id.ee/) and visiting the closest healthcare provider or pharmacy to donate a blood sample. When both the sample and consent form had been registered at EstBB, a shortened online questionnaire (Supplementary Table 1) was sent to all participants. The diagnoses and medication use modules were removed from the initial biobank questionnaire for this recruitment phase, as high-quality health records could be obtained from EHIF and NHIS^1,2^, including prescriptions that are 99.9% digital^23^.

Currently, EstBB includes 212,000 mainly European-ancestry participants (∼20% of the Estonian adult population), for whom a variety of health-related and demographic information as well as biological samples have been collected (**Figure 1B-C**). The broad age distribution allows large-scale population-based studies on a variety of health-related and molecular traits across the entire adult lifespan. Similarly to other volunteer-based biobanks, female participants are over-represented in EstBB, while the high proportion of females of reproductive age is a unique feature and enables robust investigations into women’s reproductive health conditions^24–29^.

High-coverage whole-genome sequencing (WGS) of 2,800 participants and the genotyping of all EstBB samples have been major milestones for the biobank and RbG studies in Estonia. Individuals were randomly selected for WGS by the county of birth to maximise genetic diversity. This provided the backbone for constructing a population-specific reference panel for genotype imputation^30^ and facilitated the discovery of novel population-specific genetic variants associated with diseases^19,31^ and pharmacogenetics^32^. WGS led to the identification of over 1,900 putative loss-of-function variants that have not been detected in other populations yet (compared against the Genome Aggregation Database v2.1.1 (exomes and genomes) and v3.1.2)^33^. Additionally, whole exome sequences (WES) are available for 2,500 participants, mainly constitutionally thin individuals and healthy controls^34^. Today, all participants have been genotyped using the Global Screening Array (Illumina Inc. USA), which contains >780,000 markers across the genome, and specific add-on content of 2,000 novel potentially high-impact variants identified by WGS and WES in the Estonian population. Genotype quality control follows a rigorous in-house pipeline with new releases launched promptly as new individuals are enrolled. Other layers of omics data collected over the years are summarised in **Figure 1C** (Supplementary Table 2), and a detailed overview of specific data types available at EstBB is provided below.

### Overview of data available in EstBB

The data available in EstBB encompass various *omics*-layers and their derivatives, as well as questionnaires and health-related information extracted from different national registries and more complex digital EHRs (‘deep phenotyping data’), providing an unprecedented level of detail. Estonia has been among the global leaders in digitalizing its healthcare system, having implemented several nationwide e-Health solutions that integrate data from primary and specialist care, and an infrastructure that facilitates data linking^23^. While structured information on diagnoses, prescriptions and medical procedures can be obtained via linking to EHIF and NHIS, the electronic data also include more detailed, often free-form medical records from hospitals and healthcare providers. This rapidly growing collection of records contains more than ten different types of documents dating back to 2004, marking the earliest entries in the centralized EHRs. Collaborative efforts led by STACC and the University of Tartu have driven the transformation of these data to a research-ready format. These include direct parsing, data cleaning and standardization, but also more complex natural language processing tasks for structuring the information represented in free-text format^35,36^.

#### Data derived from genotype data

##### Structural variation

CNV have been detected for the genotyped EstBB cohort using the commonly used array-based calling software PennCNV^37^. As CNV discovery is highly dependent on the array signal quality, we have performed extensive sample quality control^38^ and adding our own omics-informed quality score^39^. We retained 937,747 high-confidence deletion and 673,113 high-confidence duplication calls for 191,469 EstBB samples (on average, 4.90 deletions and 3.52 duplications per sample) (**Figure 2 A-B**). As is typical for array-based calling, the majority of CNVs are rare with only 0.49% of the genome (calculated based on GSA probe positions) having CNV overlap frequencies >0.01. In addition to the analysis of syndromic rare CNVs^17,40–43^, the EstBB CNV dataset has proven to be a valuable resource for the genome-wide assessment of rare and common CNVs associated with clinically relevant traits and diseases^38,44^.

**Figure 2.**
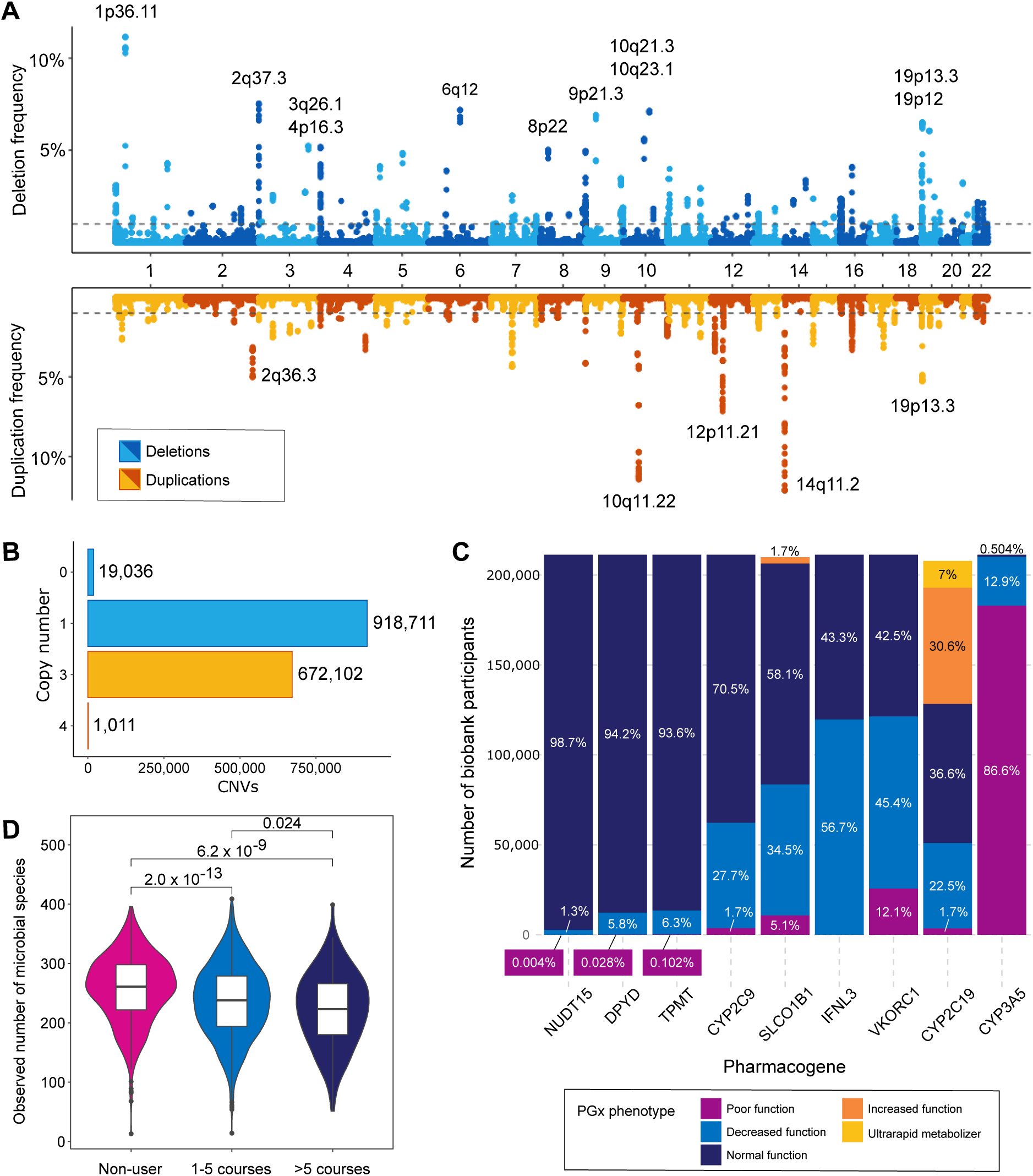
A) CNVs in the EstBB cohort. Deletion (blue) and duplication (yellow) frequencies (y-axis) at GSA probe positions (x-axis) are presented. The dashed line indicates 1% frequency. Loci with CNV frequencies >5% are labelled with cytogenic bands. B) Numbers of detected CNVs (x-axis) with copy number 0 to 4 (y-axis). C) Number of individuals (y-axis) and frequencies of assessed pharmacogenetic phenotypes of nine major pharmacogenes (x-axis). The proportions of PGx phenotypes among all analysed individuals (N=211,257) are shown with percentages on each bar. Different colours correspond to the established PGx phenotypes for each gene. D) Observed number of microbial species (y-axis) among the microbiome cohort participants who had taken 0, 1-5 or >5 courses of antibiotics within 5 years prior to microbiome sample collection. Individuals who used antibiotics within 6 months before sampling were excluded.

##### *HLA* allele imputation

To improve the discovery of variation within the human leukocyte antigen (HLA) genes with complex traits, HLA imputation was carried out based on a merged reference of EstBB WGS^30^ and Type 1 Diabetes Genetics Consortium data using the SNP2HLA tool^45^. To date, HLA imputation data are available for all EstBB participants, and this dataset has been central to characterising HLA allelic associations with penicillin allergy^46^, cervical malignancy^47^, pernicious anemia^48^, and has been used to validate associations found in exome sequencing based HLA allele analysis in the UK Biobank (UKBB) data^49^.

##### Translating genotypes into pharmacogenetic phenotypes

The Clinical Pharmacogenetics Implementation Consortium (CPIC)^50^ and the Pharmacogenomics Knowledge Base (PharmGKB)^51^ provide curated clinically relevant information for personalised drug therapy – pharmacogenetics^52^. Using these resources and the PharmCAT algorithm^53^ on phased genotype data, EstBB has established pharmacogenetic phenotypes and recommendations for drug therapy based on nine pharmacogenes for 211,257 individuals. Almost all individuals (99.99%) carry a genetic variant in at least one of the studied genes with a recommendation for dose adjustment or change in medication (**Figure 2C**, Supplementary Table 3). Star allele calls for the major drug metaboliser CYP2D6 are currently being validated based on long-read sequencing of EstBB samples to ensure the accuracy of called single nucleotide variants, structural variants, and their phase.

#### Omics data

##### Biomarker profiling

The plasma samples of EstBB participants have been profiled using several metabolomics platforms. In collaboration with Nightingale Health, nuclear magnetic resonance (NMR) was used to generate plasma metabolite profiles for all individual samples in the biobank (**Figure 1C**)^54^. The assay covers 249 biomarkers ranging from low molecular weight compounds to lipids and lipoproteins^55^. An earlier set of NMR metabolomics data for 11,000 biobank participants has been used in studies examining genetic effects on metabolic traits^56,57^ and their associations with cardiometabolic outcomes^56,58^, as well as investigating changes in metabolite levels in relation to kidney cancer^59^, telomere length^60^, and all-cause mortality^61^. Notably, four biomarkers were identified as predictive of mortality from cancer, nonvascular causes, and cardiovascular events, implying systemic interconnections among seemingly unrelated health conditions. Integration of these biomarkers into risk prediction models has resulted in improved accuracy in estimating 5-year mortality rates^61^.

EstBB has additionally generated several smaller metabolomic datasets. These include a mass spectrometry (MS) based profile assessed with the AbsoluteIDQ p150 Kit (Biocrates Life Sciences) covering 190 markers such as acyl carnitines, amino acids, glycerophospholipids, sphingolipids, and hexoses for 1,100 individuals^62^. The liquid chromatography (LC) MS-based Metabolon platform was used to profile 1,505 endogenous and exogenous metabolites for 990 participants, enabling an in-depth investigation of the metabolic risk factors associated with 14 non-communicable diseases^63^. An additional LC-MS dataset (Q Exactive, Thermo Scientific) was profiled for 580 individuals, allowing further characterisation of all-cause mortality^64^. Lastly, as part of the EXPANSE project^65^, gas chromatography MS and high-resolution LC-MS are being generated for an additional 1,000 EstBB participants.

Other datasets include clinical biochemistry measurements for 2,700 individuals (45 metabolites and parameters commonly measured at the hospitals), IgG glycosylation markers (78 in total) available for 1,055 individuals, concentrations of 341 proteins detected by the Olink Proteomics platform for 500 individuals, and 4,679 unique protein levels measured on the SomaLogic platform with SomaScan for 580 samples. The Olink dataset has been used for a thorough investigation of the effects of common, rare and structural variation on plasma protein levels^66^. DNA methylation data has been generated for 700 individuals and allowed studies on disease and age-related changes in the epigenome of leukocytes and purified CD4+, CD8+ T-cells and monocytes^67–72^. Also, gene expression profiles have been generated with stranded RNA-seq (600 individuals) and Illumina expression arrays (900 individuals) from blood and several blood cell subtypes. These data have revealed novel insights into autoimmune diseases^73^ and immune response^74^, as well as used in large-scale consortium meta-analyses, aiming to decipher the molecular mechanisms by which genomic variation can impact complex traits^75,76^.

Recent text-mining efforts of structured and unstructured data from EHRs have resulted in an additional compilation of a comprehensive dataset of biomarkers from blood and urine, continuously collected since 2004. This growing data set encompasses clinical biomarker entries, identified and categorized using LOINC codes, from hospitals and health system laboratories, as well as from written medical case reports (epicrises), with a thoroughly cleaned version currently holding 4.8M entries for EstBB participants. Among other applications, these extensive, continually updated data are used in hormonal biomarker GWAS studies^12^ and coupled with machine learning approaches, have revealed longitudinal trends in common clinical parameters indicative of future disease events, such as ischemic stroke^77^. Furthermore, extracting multiple body mass index (BMI) datapoints from various EstBB-linked records allowed researchers to determine longitudinal BMI trajectories to dissect factors driving the increased prevalence of metabolic syndrome among individuals with schizophrenia spectrum disorder^78^.

##### Microbiome data

In 2017-2019, EstBB established the Estonian Microbiome (EstMB) cohort and collected additional oral, stool and plasma samples from 2,509 EstBB participants for microbiome studies. The metagenomes of the stool samples have been characterized using shotgun metagenomic paired-end sequencing on the Illumina NovaSeq 6000 platform (1.96 ± 0.20 Gb, 15.3M ± 1.55M host-cleaned paired-end reads per sample), and a subset of 1,878 samples have additionally been sequenced by MGI technology (11.7 ± 0.20 Gb, 56.1M ± 19.4M host-cleaned paired-end reads per sample). Additionally, the participants have filled in a microbiome-related questionnaire that complements the EstBB core questionnaire data with more in-depth information on diet, lifestyle and environment. The microbiome dataset has been used to characterize the factors associated with faecal microbiome structure^79,80^ and for developing novel computational tools^81,82^. Notably, by leveraging digital drug dispensing data over a 10-year period, a study on the EstMB cohort characterised the long-term effect of antibiotics usage on the microbiome (**Figure 2D**)^79^. A follow-up study using faecal samples of EstMB participants for microbiota transplantation revealed a significant antibiotic-induced physiological effect on the gut barrier function^83^. Recently, a second sample collection for a subset of 328 participants has enabled the systematic evaluation of long-term drug effects across various drug classes in addition to antibiotics (manuscript in preparation). Finally, deep metagenomic sequencing of 1,898 faecal samples has facilitated the assembly of 84,762 metagenome-assembled genomes, including 353 (16%) previously unidentified or potentially novel species (manuscript in preparation). By providing a population reference, this dataset will serve as a valuable resource for microbiome-based association studies.

#### Overview of health and phenotype data

##### Health data from registries

The health records of EstBB participants are regularly updated by linking to the national health databases of EHIF (treatment bills and prescriptions), NHIS, the Cancer Registry, the Myocardial Infarction Registry, and two major hospitals in Estonia (North Estonia Medical Centre and Tartu University Hospital) (Supplementary Figure 1). Linkage to the Population Registry and the Causes of Death Registry provides information on deceased participants. Treatment bills and prescription data from EHIF are available for virtually all EstBB participants (>99.5%), and >97% have at least one full text record (medical case report) from NHIS (Supplementary Table 4).

Based on the latest linkage to the EHIF database in December 2023, more than half of the EstBB participants have at least one record of upper respiratory tract infections, dorsalgia (back pain), eye diseases, or COVID-19. Approximately one-third of the biobank participants have records of hypertension or dyslipidaemia, and 40,469 have been diagnosed with arrhythmias, 24,019 with *angina pectoris,* 17,888 with chronic ischemic heart disease, and 17,838 with type 2 *diabetes mellitus*. One-quarter of the participants have been diagnosed with major depressive disorder, 47,331 with anxiety disorders, and 40,861 with sleep disorders (Supplementary Table 5). As of March 2024, 5% of the biobank participants (n=10,730) have passed away, with the most common causes of death summarised in Supplementary Table 6. Additionally, EHRs allows further extraction of detailed data, such as information about medical procedures, treatments, and cancer stages.

##### Prescription data

Antibiotics are the most prescribed medications in the cohort with approximately 2 million purchases by 204,030 individuals. Amoxicillin, for example, has been prescribed at least once to 132,142 individuals (312,970 purchases). Similarly, clarithromycin has been prescribed to 115,385 individuals (272,721 purchases) (**Figure 3A**). The next most prescribed drugs include anti-inflammatory and antirheumatic products (ATC code M01) for 166,375 participants, corticosteroids as a dermatological preparation (ATC code D07) for 115,264 participants, and ophthalmological agents (ATC code S01) for 113,225 individuals. Among the most purchased medications are drugs acting on the renin-angiotensin system (ATC code C09; 2.8 million purchases), sex hormones and modulators of the genital system (ATC code G03; 2.1 million purchases), beta-blocking agents (ATC code C07; 1.8 million purchases), and anti-inflammatory and antirheumatic products (1.7 million purchases). Utilizing such data across three large-scale biobanks revealed highly polygenic architecture for lifelong medication use in cardiometabolic conditions ^5^.

**Figure 3.**
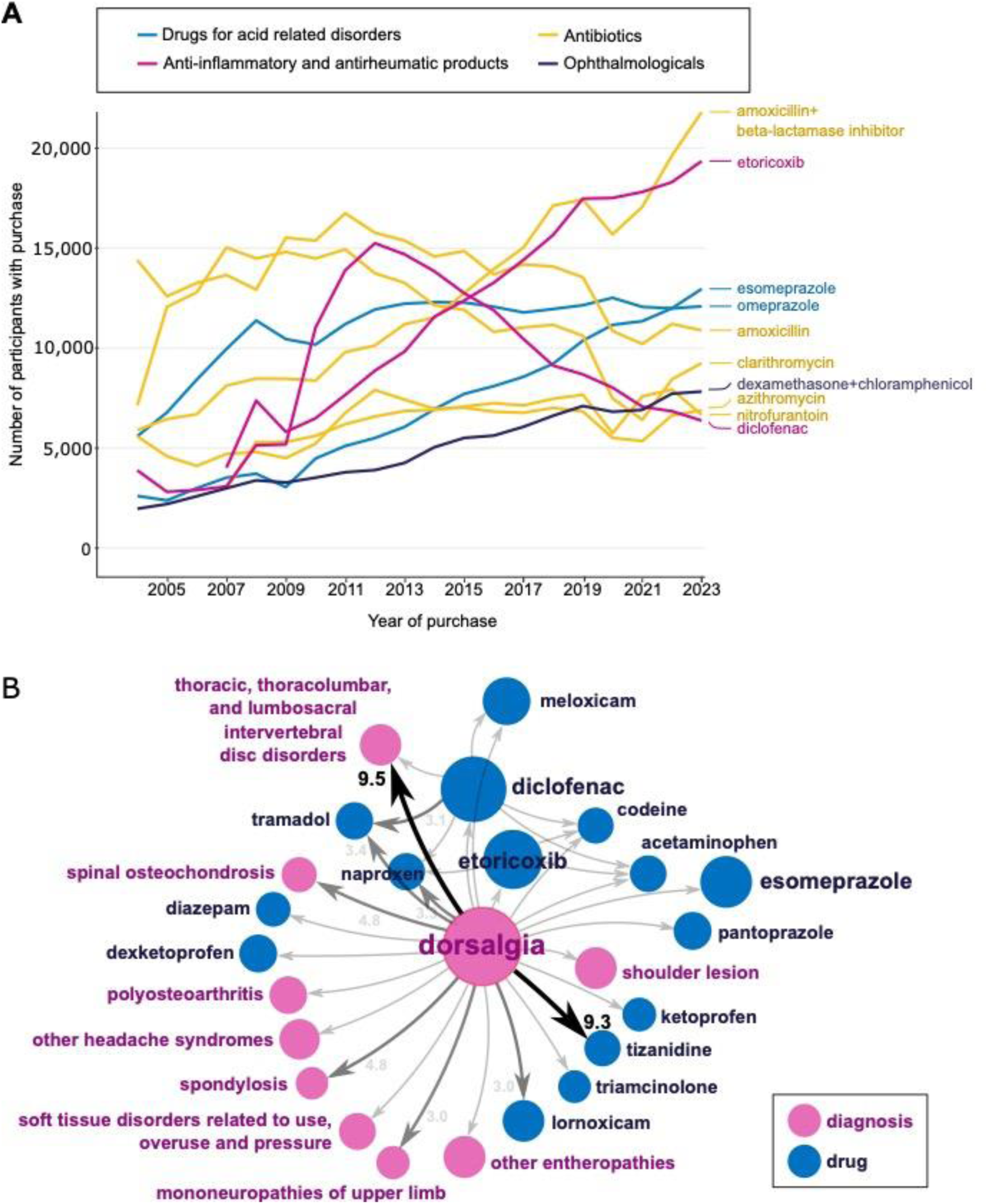
A) Number of EstBB participants with top 10 purchased medications from 2004 to 2023. Different coloured lines indicate different medication classes and reflect the number of EstBB participants who were prescribed the respective drugs each year. B) Illustration of disease and treatment trajectories detected in EstBB, exemplified by dorsalgia as a starting point. Prior dorsalgia significantly increases the relative risk of observing subsequent diagnoses (purple) and medications (blue) in the dataset. Node size indicates the number of patients, and numbers on the arrows denote the relative risk (only shown if relative risk is greater than 3).

However, the drug prescription database that covers all out-patient drug prescriptions and purchase information does not include data on drugs administered in hospitals or those sold over the counter without a prescription. While some of this information can be retrieved from medical case reports, which may provide details about active ingredients encoded as ATC codes and prescribed drug doses, these are often included in voluntary free-text sections, making complete coverage of medication data challenging.

Pharmacogenetics provides an example of the value of text extraction from EHRs particularly for studying adverse drug reactions (ADEs). This requires complex tasks involving combinations of different lexicons, linguistic rules, edit distances and regular expressions. ADEs typically involve at least two entities in the text – the drug name (or substance or other more general identifier, e.g., ‘antibiotics’) and the reaction itself. For instance, the extraction of self-reported penicillin allergies using regular expressions for both entities as well as distance rules between them led to the discovery of cases completely absent in structured EHR data. A meta-analysis of GWAS including data from EstBB, UKBB and Vanderbilt University Medical Center’s BioVU, coupled with in-depth analysis of the HLA region, led to the identification of genetic variation linked with penicillin allergy^46^. A similar approach to extract all ADEs mentions across different drug groups from free-text fields is currently under way.

Health data from registries, EHRs and the prescriptions database are harmonized to common vocabularies and made available in Observational Medical Outcomes Partnership (OMOP) common data model format^35^. The OMOP version of the data is used in the Analysis and Real World Interrogation Network (DARWIN EU) for providing real-world evidence for regulatory purposes of medicines (https://www.darwin-eu.org/). The internal event pathways within the clinical datasets have been investigated to identify the most prominent sequences between diseases and medications^84^. The analysis revealed 94 statistically significant temporal event pairs observed among at least 5% of the EstBB participants, in which the occurrence of the first event increases the risk of the second event by at least 2-fold. For example, 24 of 94 pathways begin with the diagnosis of dorsalgia **(Figure 3B).** For 38,984 people (19.6%), it is followed by the prescription of diclofenac for relieving the symptoms of back pain.

##### Questionnaire data

In addition to health records linked from the national and hospital databases and registries, multiple self-report questionnaires provide an additional layer of data not easily retrievable from EHRs, such as lifestyle and sociodemographic factors, anthropometry, dietary information, female health and medication side-effects. The first 52,000 participants recruited in 2004-2010 underwent a computer-assisted personal interview at recruitment, guided by a medical professional (GP, nurse) and documented directly as a structured electronic questionnaire (Baseline Questionnaire 1)^1^. Participants recruited from 2018 onwards have filled out an online questionnaire after recruitment (Baseline Questionnaire 2), covering similar domains as in the first phase, but with less detail. The baseline questionnaire data are currently available for approximately 159,000 individuals (Supplementary Table 1).

The first enrolment was mainly done through a network of GPs and other medical personnel in hospitals, private practices and special recruitment offices established by EstBB, the second wave involved large media campaigns with simplified procedures, requiring signing an online consent form and donating blood at the closest healthcare provider or in a pharmacy. While both recruitment phases captured individuals with similar age and sex proportions, the first phase included more non-Estonians, fewer individuals with a university degree, slightly more individuals with BMI >30, and more reporting current smoking, compared to the second wave, based on the baseline questionnaire (Supplementary Table 7). Owing to different recruitment strategies used in the two recruitments, these results reflect a possible “healthy volunteer bias” in the second recruitment wave, which is often seen in volunteer-based biobanks.

Additionally, three large online questionnaire-based data collections have recently been conducted. The mental health online survey (MHoS) “Wellbeing and Mental Health” data collection spanned from March to July 2021. The questionnaire covered self-reported current and lifetime symptom level information, assessed with brief screening instruments, on a broad range of common psychiatric disorders, their risk factors and medication effects and side effects^85^. The MHoS study had 86,000 respondents, with an overall response rate of 47%. This survey was followed by a personality survey (PS21) from November 2021 to March 2022, covering the Big Five personality domains and other personality traits (rated by participants themselves and, optionally, their close others), as well as attitudes, life satisfaction, socioeconomic characteristics, and recent life events. With 77,400 participants, the PS21 had a response rate of 42%^86^. These data allow studying the causes and consequences of mental well-being and the role of behavioural traits in overall health. More recently, a questionnaire targeting side-effects experienced from medications and vaccines (ADE-Q) was implemented in EstBB. These data were collected over six months, from April to September 2022. Over 45,000 individuals completed the questionnaire, and 31.1% reported at least one side-effect for a drug. For all online data collections, the invitations to participate were sent out to all living EstBB participants who had not opted out of getting recontacted and had a valid email address (n=185,000). Further online data collections are planned or already ongoing, providing follow-up data for previously assessed variables and enriching EstBB with new modules covering health-relevant fields of scientific interest (e.g. reproductive health, cognitive abilities, etc).

### Implementation of EstBB data: from dissecting common trait variation to improving clinical practice

The treasure-trove of genetic and phenotypic data accumulated in biobanks can be used in a myriad of ways, ranging from improved understanding of human health, behaviour, and well-being to methodological developments and clinical studies. Next, we provide examples of how EstBB data has already advanced science beyond classical genotype-phenotype association studies.

#### Common variation linked with health-related traits and methodologies developed in EstBB

EstBB has actively contributed to advancing methodologies for genetic risk prediction of common traits. These efforts range from refining the earliest approach of summing GWAS-significant genetic variants by double-weighting effect sizes of genetic variants to improve PRS performance^21^ to building a standardized framework in collaboration with five large-scale biobanks^87^. EstBB has also contributed to evaluating the predictive value of PRSs in relation to current clinical practice and among individuals of different ancestries. For instance, a metaGRS for breast cancer showed strong predictive ability in the Estonian population, with the hazard for women in the top 5% of the PRS distribution being almost three times higher compared to those close to the median. Furthermore, women in the highest five percentiles of the PRS distribution reached a cumulative risk level of 5% by age 49, more than 20 years earlier than the population average. Conversely, women below the median PRS level reached the same risk level by age 79, nearly 10 years later^20^. Additionally, combining local ancestry deconvolution and partial risk score computation provided a significant improvement in PRS predictive ability among admixed individuals and in correcting for population-based bias^88^. These efforts, among others^88,89,91^, are laying the groundwork for improving nationwide screening programs and personalizing prevention strategies.

EstBB genetic data has offered valuable insights beyond the genetic architecture of the complex diseases prevalent today, extending to their evolutionary origins^92–95^. For instance, investigating the unique combination of genetic variants inherited from diverse ancestral components has shed light on the selection patterns influencing complex traits among contemporary Europeans. By leveraging genotype and present-day phenotype data from EstBB and ancient genomic data, substantial ancestral differences were identified for several traits, including BMI, height, caffeine consumption, blood pressure, age at menarche, eye and hair colour, cholesterol levels, and sleep patterns, with evidence of positive selection for certain traits^92^. Additionally, an in-depth analysis using WGS data revealed that a sizable fraction of risk loci associated with inflammatory conditions show signatures of positive natural selection acting on the risk alleles, providing additional evidence for pleiotropic effects of these alleles. Combining association statistics with information about the strength of selection allowed to further finemapping of some of the potentially causal variants which can be targets for future functional experiments^95^. EstBB data has been valuable in reconstructing the population history of Estonia and beyond. Sharing patterns of Identity by decent (IBD) segments among 2500 high coverage EstBB genomes revealed detailed genetic structure in Estonia mirroring geography. Reconstructions of regional dynamics of effective population size over past millennia correlate with known historic population catastrophes like wars, famine and epidemics^96^. Combining modern data with ancient DNA has revealed that the contemporary genetic structure in Estonia goes back to the Iron Age^97^.

Besides tools developed for common variant discovery^98–103^, in-house pipelines are in place for both phenome-wide associations studies (PheWAS) and phenotype-phenome wide association studies (PhePheWAS), offering insights into pleiotropic mechanisms and phenotypic comorbidities. These methods were employed to explore the comorbidities of attention-deficit hyperactivity disorder (ADHD) by assessing the associations of its genetic liability with other medical conditions in EHRs. The findings indicated significant associations with chronic obstructive pulmonary disease, obesity, and type 2 *diabetes mellitus* in undiagnosed individuals^104^, underscoring the importance of early intervention and comprehensive management strategies.

#### Re-contacting biobank participants based on genetic findings

Population or volunteer-based biobanks have opened an opportunity for research on communicating genomic risk information to research participants. This includes both symptomatic and apparently healthy individuals carrying established pathogenic or likely pathogenic genetic and structural variants who could benefit from awareness of the risk of their genetic findings. To assess the feasibility and impact of such RbG interventions, EstBB has initiated and completed several projects inviting individuals enrolled at the biobank to participate in studies where they were offered individual genetic research results (**Figure 4**). Invitations sent to the EstBB participants do not contain any information on high-risk genetic findings; instead, individuals must first sign an informed consent, indicating their willingness to participate in a study where they are offered their genetic results and counselling. That is, information is only disclosed to participants after they have provided written consent, and for high impact genetic variants (such as *BRCA1/2* variants, CNVs), a second blood sample is also taken for validation, before disclosing results.

**Figure 4.**
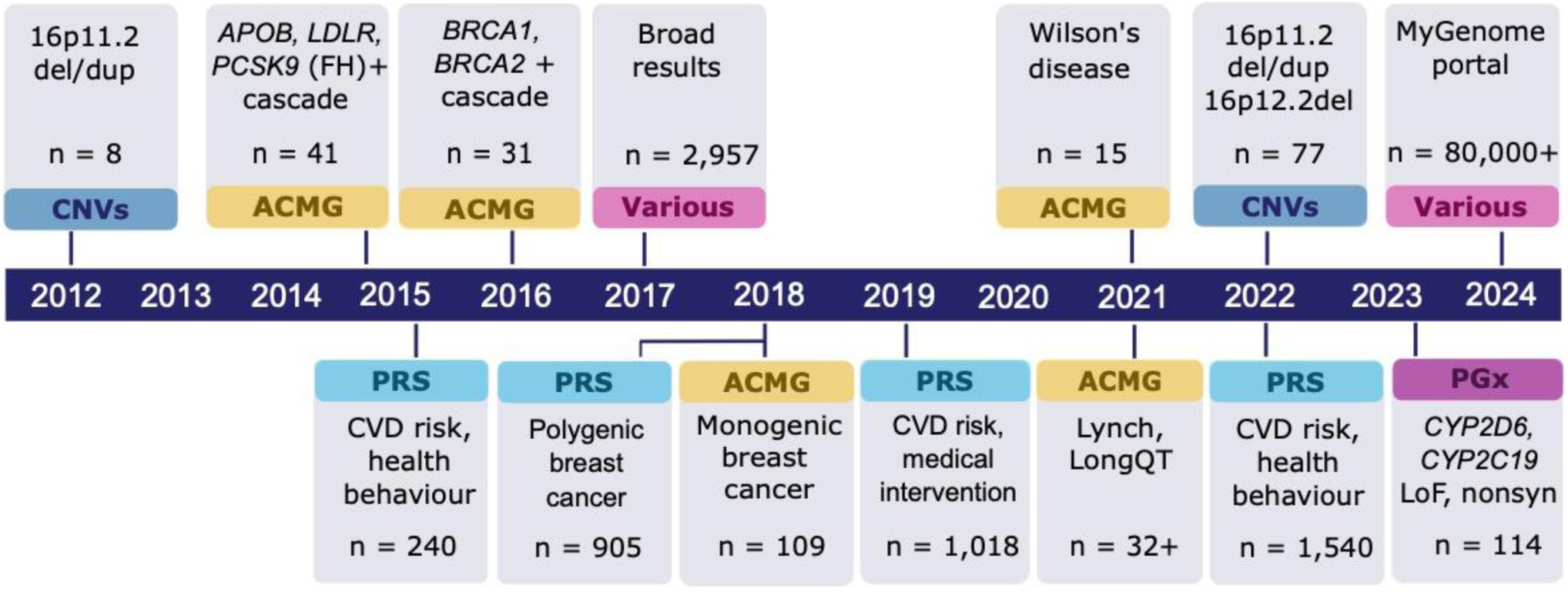
Timeline depicting recall by genotype studies carried out at EstBB. Above the timeline – return of results directly from the biobank (as a starting point); below the timeline – return of results provided in a clinical setting. The recall studies are based on CNVs – copy number variants; ACMG – American College of Medical Genetics list of genes where incidental findings should be reported; PRS – polygenic risk scores for CVD (cardiovascular disease) or breast cancer; PGx – pharmacogenetics – novel predicted LoF (loss-of-function) or nonsyn (non-synonymous) variants in CYP2D6 or CYP2C19. N indicate number of individuals with high risk PRS or variant carriers that participated in the study; FH – familial hypercholesterolemia; ‘+’ indicate on-going studies.

The recall of individuals with familial hypercholesterolemia (FH)-associated variants in the *LDLR*, *APOB*, or *PCSK9* genes was based on WES or WGS data^19^. In cascade screening involving first and second-degree relatives, 64 individuals participated, resulting in a study group of 41 carriers altogether. At the beginning of the study, 51% of FH-associated variant carriers had nonspecific hypercholesterolemia diagnosed and 7% had a clinical FH diagnosis. During the study, 51% participants were reclassified from having nonspecific hypercholesterolemia to FH, and 32% were newly diagnosed with FH, demonstrating that FH is not always recognised in the population and a RbG approach is feasible. A follow-up study conducted five years later revealed that recalled participants were more effectively engaged in the medical system and were on treatment compared to non-recalled EstBB participants carrying the same genetic variants identified in microarray data ^105^. A similar RbG approach was conducted for Wilson’s disease (pathogenic variant carriers in *ATP7B* gene) highlighting a striking 5-fold enrichment of the most prevalent causal variant in Europe (p.His1069Gln) among EstBB participants compared to other populations. This study similarly uncovered previously undiagnosed Wilson’s disease cases with mild to severe neurological symptoms detected in 87% of the individuals and biochemical alterations in all individuals^106^.

To identify individuals with elevated risk of familial breast and ovarian cancer (HBOC), carriers of 17 likely pathogenic or known pathogenic coding variants in *BRCA1 or BRCA2* from a sub-cohort of 17,679 EstBB participants were identified^18^. Both genetic counselling and a personalized surveillance plan prepared in collaboration with oncologists were provided to individuals. In a succeeding study, biobank participants with high PRS and/or monogenic variants for breast cancer were invited for an oncology visit^107^. In total, 109 women with monogenic findings participated in the study, of whom six participants received a breast cancer diagnosis and five of them before the age of 50 i.e., below the national screening program entry age. Family history was indicative of a risk variant for less than 50% of the cases, i.e., without genetic screening, over half of the subjects would not have been eligible for genetic testing according to the current criteria in Estonia^107^. The study also included 905 female participants with high PRS for breast cancer, which resulted in the identification of ten new breast cancer cases at early stages (manuscript in preparation). In both the monogenic and PRS arms of assessing breast cancer genetic risk, 98% and 100% of the participants, respectively, found the disclosure of genetic risk valuable and appreciated being contacted.

A randomized controlled trial of the effectiveness and feasibility of using PRS for the prevention of cardiovascular disease was carried out among middle-aged subjects whose PRS for coronary artery disease was in the top quintile^108^. The participants were randomized into an intervention group that received counselling about their risk (n=507), and a control group that only received the same intervention at the end of the study period (n=511). Participants in the intervention group had a significantly higher probability of initiating statin treatment than controls. Their LDL-cholesterol levels decreased significantly by the end of the study and were significantly lower than in the control group. Notably, 98.4% of the participating GPs expressed interest in integrating genetic risk assessment into their daily practice.

In all projects where genetic risks are communicated to the EstBB participants, feedback is collected to understand how participants perceive genetic information in general, as well as the impact of potentially unexpected information regarding increased genetic risk. Notably, participants tend to report positive feelings after receiving results, even when receiving high-risk information^18,109^ and, as such, provide crucial input for the broader implementation of the communication of individual research results to biobank participants.

The groundwork for the large-scale implementation of communicating individual genetic results to unselected EstBB participants was laid by a pilot study conducted in 2017-2018 which relied on the face-to-face delivery of results^109^. It covered a wide range of results, including genetic risk for type 2 *diabetes mellitus* and coronary artery disease, and recommendations of how this risk can be modified by improving lifestyle. Additionally, pharmacogenetic information for 28 medications, and carrier status for genetic variants, which may not pose an immediate threat to the individual but could impact offspring, were included. The feedback collected from the pilot study shed light on the reactions and sentiments of the EstBB participants specifically and potential responses to genetic risk communication more generally, and thus paved the way for launching the communication of results to all EstBB participants within the MyGenome Portal. It was piloted with the first 10,000 participants in early 2024 and opened to all biobank participants in June 2024. During the first two weeks, over 80,000 participants logged in to the portal and signed the dynamic consent where they can select which areas they would like to receive results in^110^.

In summary, these studies illustrate that 1) many individuals carrying clinically significant genetic variants currently go undetected in the medical system; 2) Recall by genotype for disease risk communication is feasible at least at a medium scale and appreciated by the participants of a volunteer-based biobank; 3) integration of clinical examination and counselling for a personal disease prevention or management plan can be crucial for the success of such interventions; and 4) disclosing genetic risk information for clinically significant actionable conditions is highly valued and does not necessarily increase anxiety among biobank participants consenting to participate in such studies.

## Conclusions

EstBB, having recently celebrated its 25^th^ anniversary, has now come of age. Here, we provided an updated overview of EstBB, highlighting its uniqueness and the depth and breadth of available data for studying the effects of genetic variation and quantitative phenotypes on health-related traits while keeping the biobank participants engaged through various studies. Based on regular surveys in the general population, ca 80% are aware and 70% supportive of the activities of the biobank (Supplementary Figure 2). Based on the data presented above, the main unique characteristics of EstBB are summarised below.

The first distinguishing feature is the high-quality, detailed, and multilayered phenotype data sourced from all levels of the healthcare system, ranging from primary care to in-patient hospital and specialist records for all age groups in EstBB. This allows not only large-scale studies across the entire adult lifespan, but also offers the appropriate framework for developing personalized medicine solutions.

Second, while in the *omics* world, power is in the numbers, with larger datasets or multi-centre collaborations and meta-analyses holding the key to success, specific populations with unique genetic makeup (such as the FinnGen study or EstBB) can provide leverage in unravelling associations that might remain undetected in other populations^111^. From this population genetics standpoint, EstBB represents a ‘*genetic bridge*‘ connecting the bottlenecked Finnish population and the Northern and Central European populations^111^. This can provide valuable insight into biological mechanisms and inform drug development.

Third, considering the population size of Estonia (approximately 1.3M), EstBB includes roughly 20% of the entire Estonian adult population, and is therefore an excellent foundation for nation-wide implementation of personalised medicine. The large proportion of related individuals (including nuclear families) open further avenues for family-based studies, setting EstBB apart from many other large-scale population-based biobanks. For example, 90% of EstBB participants have third degree or closer relatives in the same biobank, while in UKBB and FinnGen, the numbers are 30% and 74%, respectively^111,112^.

Fourth, the possibility to re-contact the participants offers opportunities for RbG approaches and collection of additional data layers (e.g., in-depth dissection of the microbiome or targeted questionnaire-based studies), as well as evaluate participants’ attitudes and reactions to receiving information about their genetic predisposition. The biobank has also established a framework for transferring high-risk genetic information to facilitate clinical interventions for participants. This work will be further expanded with the introduction of a national precision prevention service for breast cancer and pharmacogenetics services in 2025 by the Estonian Health Insurance Fund.

These strengths combined with the data layers described above, make EstBB a valuable resource for scientific discoveries, validating methods and algorithms, and replicating research findings. We expect more exciting discoveries in the future. For instance, long-read whole-genome sequencing of 10,000 samples is underway with PacBio HiFi technology, allowing to enhance the resolution of the population-specific reference panel, more accurate haplotype phasing, and resolving complex structural genomic regions crucial for calling pharmacogenomic variation. Through the collaborative EXPANSE project^65^ exposome data is being modelled and will be integrated as a new data layer into EstBB. The exposome data consists of air quality^113^, daily temperatures^114^, and additional variables such as vegetation indices, and distance to blue and green space. This opens avenues to exploring environmental influences on disease risk.

Comprehensive health and omics data of individuals spanning a broad age distribution, coupled with the possibility of recontacting participants for further studies based on specific findings sets EstBB in a unique position for discoveries in human genomics and clinical studies that bridge the gap between the development of robust risk models and their implementation in clinical practice. Advancing the science behind and working towards the implementation of personalized medicine has been at the core of the activities conducted at the biobank, which has now reached maturity.

## How to access the data?

Pseudonymised data and/or biological samples can be accessed for research and development purposes in accordance with the Estonian Human Genome Research Act (https://www.riigiteataja.ee/en/eli/ee/531102013003/consolide/current). To access data, the research proposal must be approved by the Scientific Advisory Committee of the Estonian Biobank as well as by the Estonian Committee on Bioethics and Human Research. For more details on data access and relevant documents, please see https://genomics.ut.ee/en/content/estonian-biobank#dataaccess.

## Data Availability

This is a overview paper on the Estonian Biobank. All data presented in the manuscript can be requested at

https://genomics.ut.ee/en/content/estonian-biobank

## Ethical Standards

The activities of the Estonian Biobank are regulated by the Human Genes Research Act, which was adopted in 2000 specifically for the operations of the Estonian Biobank. In this study, analysis of individual level data of the Estonian Biobank was carried out under ethical approval nr 1.1-12/624 and its extensions from the Estonian Committee on Bioethics and Human Research (Estonian Ministry of Social Affairs). All actions are in concert with the General Data Protection Regulation of the European Union.

## Acknowledgements

We would like to thank all the current and former staff at the Estonian Biobank for their dedicated work in building the biobank and making the data available for scientists: Ene Mölder, Atso Jõks, Kairit Mikkel, Mari-Liis Tammesoo, Krista-Roberta Saviauk, Mairo Puusepp, Innar Hallik, Roger Kikkas, Lars Johannes Sissas, Marielle Lepson, Viljo Soo, Heidi Saulep, Eva Mekk, Kristina Lokotar, Martin Tootsi, Helja Kabral, Krista Kruuv-Käo, Jaanus Pikani; and supporting staff at the Institute of Genomics: Krista Liiv, Merilin Raud, Tuuli Reisberg, Paula Ann Kivistik, Kadri Raav, Monika Nõmm, Merit Kreitsberg, Kaisa Kuus, Oliivika Zeiger, Signi Savi, Kadri Maal, Riina Raudne, Kristjan Karron; the support team at the High-Performance Computing Center of University of Tartu: Ivar Koppel, Sander Kuusemets, Ulvi Gerst Talas, Ott Eric Oopkaup, Sander Kütisaar, and Vladislav Tuzov. We would also like to acknowledge the continuous funding and support from the Ministry of Social Affairs. This work was supported by the Estonian Ministry of Education and Research (Teaming for Excellence and Centres of Excellence grant TK214 Centre of Excellence for Personalised Medicine), the European Union through the European Regional Development Fund (Project No. 2021-2027.1.01.24-0444), the Estonian Ministry of Education and Research, the European Union’s Horizon Europe research and innovation programme under grant agreement No 101060011. Views and opinions expressed are however those of the author(s) only and do not necessarily reflect those of the European Union or European Research Executive Agency. Neither the European Union nor the granting authority can be held responsible for them.

## Author contributions

LM, MA, TL, SvL, SR, and TH wrote the initial manuscript; AM, HA, AA, KrM, SS, LM, TE managed data and sample collection for the biobank; MJ, SR, KrK, MP, KaK, MA, DS, OA, LM, and LL made the figures and tables of the manuscript; all authors contributed to writing the manuscript and revising the manuscript. All authors have read and approved the final version of the manuscript.

