## Supplementary Information for "From Biobanking to Personalized Medicine: the journey of the Estonian Biobank"

Affiliations:
[1] Estonian Genome Centre, Institute of Genomics, University of Tartu, Estonia
[2] Estonian Biobank, Institute of Genomics, University of Tartu, Estonia
[3] Institute of Computer Science, University of Tartu, Estonia
[4] STACC, Estonia
[5] Institute of Mathematics and Statistics, University of Tartu, Estonia
[6] Estonian Biocentre, Institute of Genomics, University of Tartu, Estonia
[7] Institute of Psychology, University of Tartu, Estonia
[8] Centre for Genomics, Evolution and Medicine, Institute of Genomics, University of Tartu, Estonia

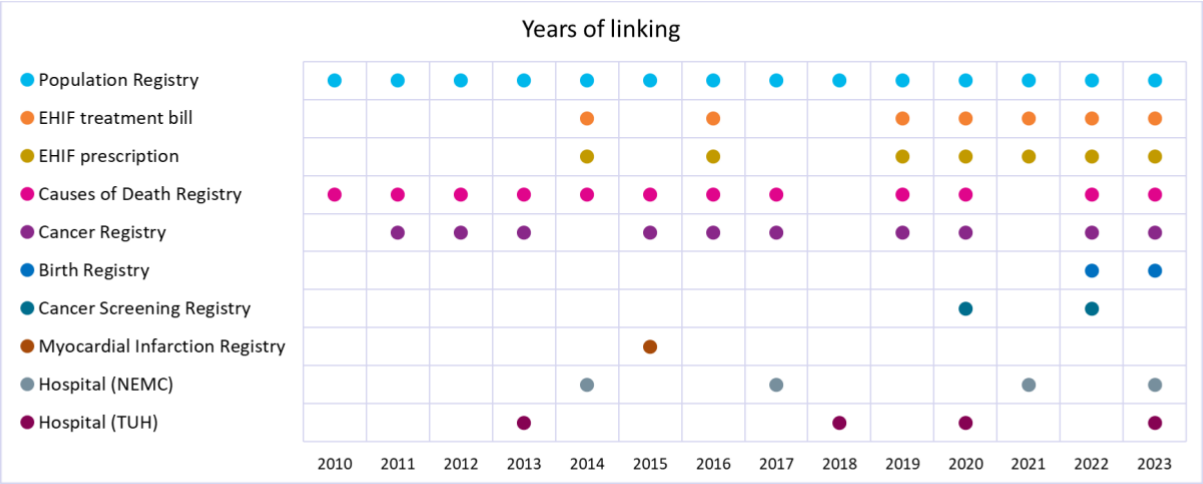


**Supplementary Figure 1.** Overview of different registries and data sources, and the frequency of data linking in EstBB. Different colored dots represent different sources (see legend on the left). EHIF – Estonian Health Insurance Fund, NEMC – North Estonian Medical Centre, TUH – Tartu University Hospital.

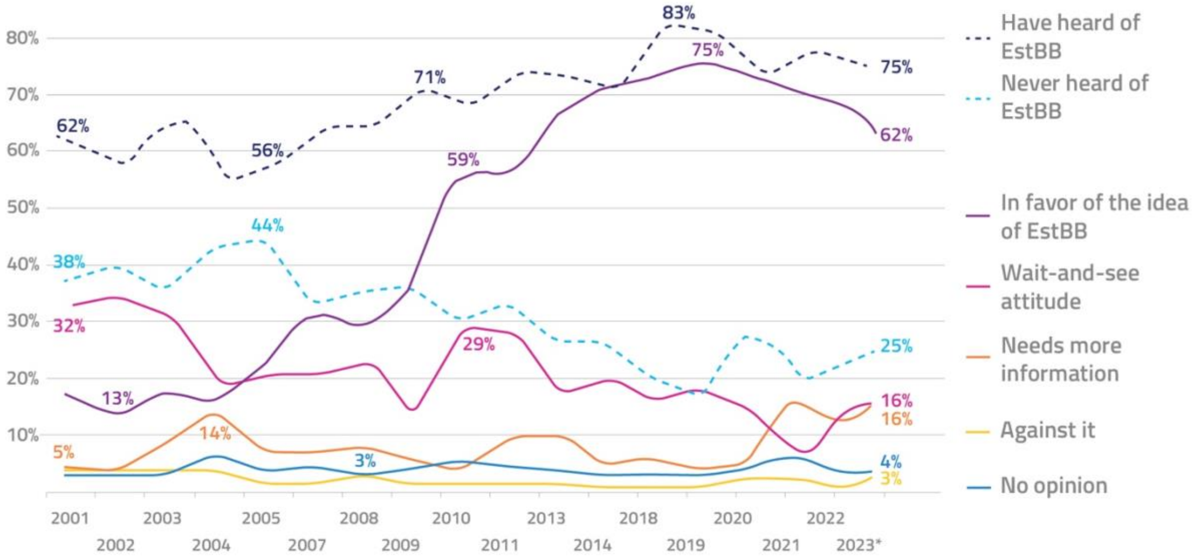


**Supplementary Figure 2.** Public awareness (dashed line) and attitudes (solid line) towards EstBB in the Estonian population. Each year, ~1000 Estonian residents between the ages of 15 and 74 years were surveyed by a polling agency (TNS Emor). The sample was composed to be proportional to the population structure with respect to age, sex, region, and nationality. *) The 2023 survey was finalized in January 2024.

**Supplementary Table 1.** Overview of the EstBB questionnaire-based data collections.

| Questionnaire | Modules | Year | N of respondents | Questionnaire link |
|---|---|---|---|---|
| Baseline questionnaire 1 | • Sociodemographics<br>• Alcohol, tobacco and other substance use<br>• Physical activity<br>• Anthropometry (weight, height, waist and hip circumference) and objective measurements (blood pressure, pulse rate) *<br>• Sleep<br>• General health<br>• Medical history<br>• Female health<br>• Medical family history<br>• Dietary habits<br>• Medication use and side effects | 2004-2010 | 52,000 | |
| Baseline questionnaire 2 | • Sociodemographics<br>• Alcohol, tobacco and other substance use<br>• Physical activity<br>• Anthropometry (weight, height, waist and hip circumference) **<br>• Sleep<br>• General health<br>• Female health<br>• Dietary habits | 2018-2023 | 111,000 | |
| Mental Health online Survey (MHoS) | • Current and/or lifetime symptoms of common psychiatric disorders (depression, bipolar disorder, attention-deficit/hyperactivity disorder, eating disorders, generalized anxiety disorder, psychotic experiences, post-traumatic stress disorder, suicidal ideation and attempts, substance abuse, problematic gambling)<br>• Lifestyle (substance use, physical activity, screen time)<br>• Childhood psychosocial environment<br>• Stressful life events<br>• Social support<br>• Psychiatric medication effects and side effects | March-July 2021 | 86,000 | https://doi.org/10.1093/ije/dyae017 |
| Adverse events from medicines and vaccines (ADE-Q) | • Adverse events from medications<br>• Adverse events from vaccines<br>• Specification of medication and adverse event | April-September 2022 | 45,000 | |
| Personality | Estonian NEO-PI-3 | 2008-2015 | 2,000 | https://osf.io/97pvz/ |
| | One Hundred Nuances of Personality (100NP) | November 2021 – March 2022 | 77,000 | https://osf.io/97pvz/ |

* Anthropometry and objective measurements were determined by medical professionals.
** Anthropometry was filled by participants.

**Supplementary Table 2**. Omics datasets EstBB.

| **OMICS profiling** | **N** |
|---|---|
| Genome-wide genotyping arrays | 212,000 |
| Metabolomics (NMR) | 200,000 |
| Whole genome sequencing | 2,800 |
| Whole exome sequencing | 2,500 |
| Microbiome oral, stool (gut metagenomics) | 2,500 |
| Telomere length | 5,200 |
| Clinical biochemistry | 2,700 |
| Metabolomics (MS/MS) | 1,100 |
| Metabolomics (LC-MS and Metabolon) | 1,600 |
| Genome-wide gene expression arrays | 900 |
| IgG glycosylation | 1,000 |
| Genome-wide methylation arrays | 700 |
| mRNA sequencing | 600 |
| Proteomics (SomaLogic) | 600 |
| Proteomics (Olink) | 500 |

**Supplementary Table 3.** Frequencies of evaluated PGx phenotypes (and activity scores where available) in EstBB. The corresponding average frequencies for the European population are provided primarily based on PharmGKB gene-based frequency tables. A phenotype with a number (e.g. Intermediate Metabolizer 1.0) indicates the estimated activity score of this gene.

| Gene | PGx phenotype | N in EstBB | EstBB frequency | European frequency |
|---|---|---|---|---|
| CYP2C19 | Intermediate Metabolizer | 47,518 | 22.5% | 26.1% |
| CYP2C19 | Normal Metabolizer | 77,260 | 36.6% | 39.6% |
| CYP2C19 | Poor Metabolizer | 3,526 | 1.7% | 2.4% |
| CYP2C19 | Rapid Metabolizer | 64,606 | 30.6% | 27.1% |
| CYP2C19 | Ultrarapid Metabolizer | 14,876 | 7.0% | 4.6% |
| CYP2C9 | Intermediate Metabolizer 1.0 | 26,057 | 12.3% | 13.8% |
| CYP2C9 | Intermediate Metabolizer 1.5 | 32,543 | 15.4% | 20.8% |
| CYP2C9 | Normal Metabolizer 2.0 | 148,967 | 70.5% | 62.8% |
| CYP2C9 | Poor Metabolizer 0.0 | 1,014 | 0.480% | 0.6% |
| CYP2C9 | Poor Metabolizer 0.5 | 2,676 | 1.3% | 2.0% |
| CYP3A5 | Intermediate Metabolizer | 27,321 | 12.9% | 13.7% |
| CYP3A5 | Normal Metabolizer | 1,064 | 0.504% | 0.5% |
| CYP3A5 | Poor Metabolizer | 182,872 | 86.6% | 85.7% |
| DPYD | Intermediate Metabolizer | 12,275 | 5.8% | **3–7% |
| DPYD | Poor Metabolizer | 59 | 0.028% | **0.3% |
| IFNL3 | Favorable Response Genotype | 91,490 | 43.3% | *38.0% |
| IFNL3 | Unfavorable Response Genotype | 119,767 | 56.7% | *63.0% |
| NUDT15 | Intermediate Metabolizer | 2,722 | 1.3% | 0.8% |
| NUDT15 | Normal Metabolizer | 208,527 | 98.7% | 98.6% |
| NUDT15 | Poor Metabolizer | 8 | 0.004% | 0.001% |
| SLCO1B1 | Decreased Function | 72,912 | 34.5% | 28.3% |
| SLCO1B1 | Increased Function | 3,504 | 1.7% | 3.0% |
| SLCO1B1 | Normal Function | 122,698 | 58.1% | 65.6% |
| SLCO1B1 | Poor Function | 10,748 | 5.1% | 2.9% |
| TPMT | Intermediate Metabolizer | 13,297 | 6.3% | 8.4% |
| TPMT | Normal Metabolizer | 197,744 | 93.6% | 90.9% |
| TPMT | Poor Metabolizer | 215 | 0.1% | 0.2% |
| VKORC1 | Normal Metabolizer | 89,818 | 42.5% | 58.7% |
| VKORC1 | Intermediate Metabolizer | 95,805 | 45.4% | *41.3% |
| VKORC1 | Poor Metabolizer | 25,634 | 12.1% | |
| All genes | At least 1 non-normal PGx Phenotype | 211,241 | 99.99% | NA |

*Based on the 1 variant carrier frequency in CPIC publication. **Based on the frequencies within the CPIC guideline publication.

**Supplementary Table 4.** Overview of data obtained via linking with national registries. Average rows per participant is calculated for the subset of participants who have data from the respective source (as indicated in the "% of all participants column").

| Source | Participants | First record date | Last record date | % of all participants | Average rows per participant |
|---|---|---|---|---|---|
| Baseline questionnaire | 158,705 | 2002/10 | 2024/04 | 74.8% | 1.05 |
| *Self-reported diagnoses* | 50,809 | 1920/01 | 2018/01 | 23.9% | 10.85 |
| EHIF treatment bill | 211,767 | 2001/08 | 2023/12 | 99.8% | 121.24 |
| *EHIF diagnoses* | 211,767 | 2001/08 | 2023/12 | 99.8% | 182.02 |
| EHIF prescriptions | 211,371 | 2004/01 | 2023/12 | 99.6% | 150.21 |
| eHealth medical case report | 206,152 | 1998/01 | 2022/12 | 97.1% | 38.16 |
| *Diagnoses from medical case report* | 206,109 | 1970/01 | 2022/12 | 97.1% | 64.32 |
| Cancer events | 17,465 | 1955/08 | 2021/12 | 8.2% | 1.17 |
| Cancer screening events | 114,304 | 2015/01 | 2021/12 | 53.9% | 2.64 |
| Medical Birth Registry | 62,468 | 1992/01 | 2023/01 | 45% (of females) | 1.92 |
| Death certificates | 10,723 | 2003/05 | 2024/04 | 5% | 1 |
| NEMC medical cases | 114,098 | 1993/02 | 2020/11 | 53.7% | 10.82 |
| *NEMC diagnoses* | 108,748 | 1993/02 | 2020/11 | 51.2% | 14.99 |
| TUH medical case | 136,301 | 1999/12 | 2023/09 | 64.2% | 16.89 |
| *TUH diagnoses* | 118,879 | 1999/12 | 2023/09 | 56% | 27.02 |

EHIF – Estonian Health Insurance Fund, NEMC – North Estonian Medical Centre, TUH – Tartu University Hospital.

**Supplementary Table 5.** Thirty most common EHIF diagnoses present in EstBB.

| ICD-10 main code | ICD-10 main diagnosis | N of EstBB participants | % of EstBB participants with diagnosis |
|---|---|---|---|
| J06 | Acute upper respiratory infections of multiple and unspecified sites | 166,111 | 78.3% |
| M54 | Dorsalgia | 137,896 | 65.0% |
| U07.1-U07.2 | Emergency use for COVID-19 | 122,683 | 57.8% |
| H52 | Disorders of refraction and accommodation | 112,607 | 53.1% |
| B34 | Viral infection of unspecified site | 99,975 | 47.1% |
| J20 | Acute bronchitis | 92,492 | 43.6% |
| J02 | Acute pharyngitis | 89,552 | 42.2% |
| M25 | Other joint disorders, not elsewhere classified | 87,882 | 41.4% |
| J01 | Acute sinusitis | 79,774 | 37.6% |
| R10 | Abdominal and pelvic pain | 78,573 | 37.0% |
| U11 | Need for immunization against COVID-19 | 77,848 | 36.7% |
| M79 | Other soft tissue disorders, not elsewhere classified | 76,336 | 36.0% |
| H10 | Conjunctivitis | 75,679 | 35.7% |
| N30 | Cystitis | 73,001 | 34.4% |
| J00 | Acute nasopharyngitis [common cold] | 70,577 | 33.3% |
| I10 | Essential (primary) hypertension | 68,079 | 32.1% |
| E78 | Disorders of lipoprotein metabolism and other lipidaemias | 66,737 | 31.4% |
| J04 | Acute laryngitis and tracheitis | 66,430 | 31.3% |
| J30 | Vasomotor and allergic rhinitis | 64,724 | 30.5% |
| B37 | Candidiasis | 62,396 | 29.4% |
| K29 | Gastritis and duodenitis | 61,985 | 29.2% |
| J03 | Acute tonsillitis | 61,786 | 29.1% |
| J35 | Chronic diseases of tonsils and adenoids | 58,845 | 27.7% |
| K21 | Gastro-oesophageal reflux disease | 57,014 | 26.9% |
| D22 | Melanocytic naevinaevus | 55,938 | 26.4% |
| N76 | Other inflammation of vagina and vulva | 55,814 | 26.3% |
| L23 | Allergic contact dermatitis | 53,668 | 25.3% |
| F32 | Depressive episode | 53,428 | 25.2% |
| B35 | Dermatophytosis | 52,175 | 24.6% |
| K04 | Diseases of pulp and periapical tissues | 52,148 | 24.6% |

**Supplementary Table 6.** Twenty most common causes of death among EstBB participants.

| ICD10 code | Diagnosis | N |
|---|---|---|
| I25 | Chronic ischaemic heart disease | 1,310 |
| I11 | Hypertensive heart disease | 1,221 |
| C34 | Malignant neoplasm of bronchus and lung | 440 |
| I63 | Cerebral infarction | 360 |
| I13 | Hypertensive heart and renal disease | 350 |
| I21 | Acute myocardial infarction | 289 |
| C25 | Malignant neoplasm of pancreas | 282 |
| C16 | Malignant neoplasm of stomach | 225 |
| C50 | Malignant neoplasm of breast | 218 |
| C18 | Malignant neoplasm of colon | 209 |
| U07.1-U07.2 | Emergency use for COVID-19 | 193 |
| C61 | Malignant neoplasm of prostate | 189 |
| E11 | Type 2 diabetes mellitus | 157 |
| I50 | Heart failure | 152 |
| K70 | Alcoholic liver disease | 152 |
| C71 | Malignant neoplasm of brain | 133 |
| J44 | Other chronic obstructive pulmonary disease | 128 |
| I42 | Cardiomyopathy | 124 |
| X70 | Intentional self-harm by hanging, strangulation, and suffocation | 113 |
| C22 | Malignant neoplasm of liver and intrahepatic bile ducts | 108 |

**Supplementary Table 7.** Characteristics of EstBB participants based on two recruitment waves.

| Characteristic | Baseline Questionnaire 1 | Baseline Questionnaire 2 |
|---|---|---|
| Mean age (range) | 44.3 (18-103) | 42.6 (18-105) |
| Sex | 65.6% women | 67.9% women |
| **Nationality** | | |
| *Estonian* | 81.5% | 93.7% |
| *Russian* | 15.3% | 4.8% |
| *Other* | 3.2% | 1.5% |
| **Education** | | |
| *Primary or basic* | 17.9% | 4.7% |
| *Secondary or secondary vocational* | 57.4% | 44.4% |
| *University degree* | 24.7% | 50.9% |
| **BMI categories (%)** | | |
| *<18.5* | 2.4% | 2.2% |
| *[18.5–25)* | 44.2% | 48.5% |
| *[25.0–30)* | 31.2% | 31.2% |
| *≥30.0* | 22.2% | 18.1% |
| **Smoking status** | | |
| *Current* | 28.6% | 14.4% |
| *Former* | 13.7% | 31.4% |
| *Never* | 57.7% | 54.2% |
